# Whole-body aging mediates the association between socioeconomic status disparities and cognition and verbal fluency among U.S. older adults

**DOI:** 10.1101/2025.06.16.25329679

**Authors:** Xiaoqin Yan, Xin Le, Shengling Wang

## Abstract

**Background:** Cognitive decline is a worldwide public health issue among older populations. Socioeconomic status disparities in income, education, and social status are linked to cognitive function and verbal fluency. Although the etiology of cognitive decline remains unclear, social and biological factors are increasingly recognized as significant risk contributors. This study aims to investigate the potential associations between socioeconomic status and cognitive functions and the mediated effect of whole-body aging.

**Methods:** Data on socioeconomic status, whole-bodying aging, cognitive functions, and verbal fluency were collected from 2321 participants aged over 60 years based on the National Health and Nutrition Examination Survey 2011-2014. Independent t-tests, one-way ANOVA tests, Spearman correlation, and multiple logistic regression were used to explore the associations of socioeconomic status, whole-body aging, and cognitive functions, respectively. Mediation analyses were used to explore the mediated effects of whole-body aging on the associations between socioeconomic status and cognitive functions.

**Results:** In the multiple regression analysis models, socioeconomic status and whole-body aging were associated with cognitive function and verbal fluency (all *p* <0.05). Mediation analyses indicated that the associations between socioeconomic status and cognitive function were parallelly mediated by whole-body aging, with the proportion of mediation ranging from 3.21% to 7.65% (all *p*□<□0.05).

**Conclusion:** These findings suggested that socioeconomic status disparities increased the impairment of immediate and delayed learning ability, sustained attention, processing speed, working memory, and verbal fluency, which was possibly and partially mediated by whole-body aging.

**Graphic Abstract:** 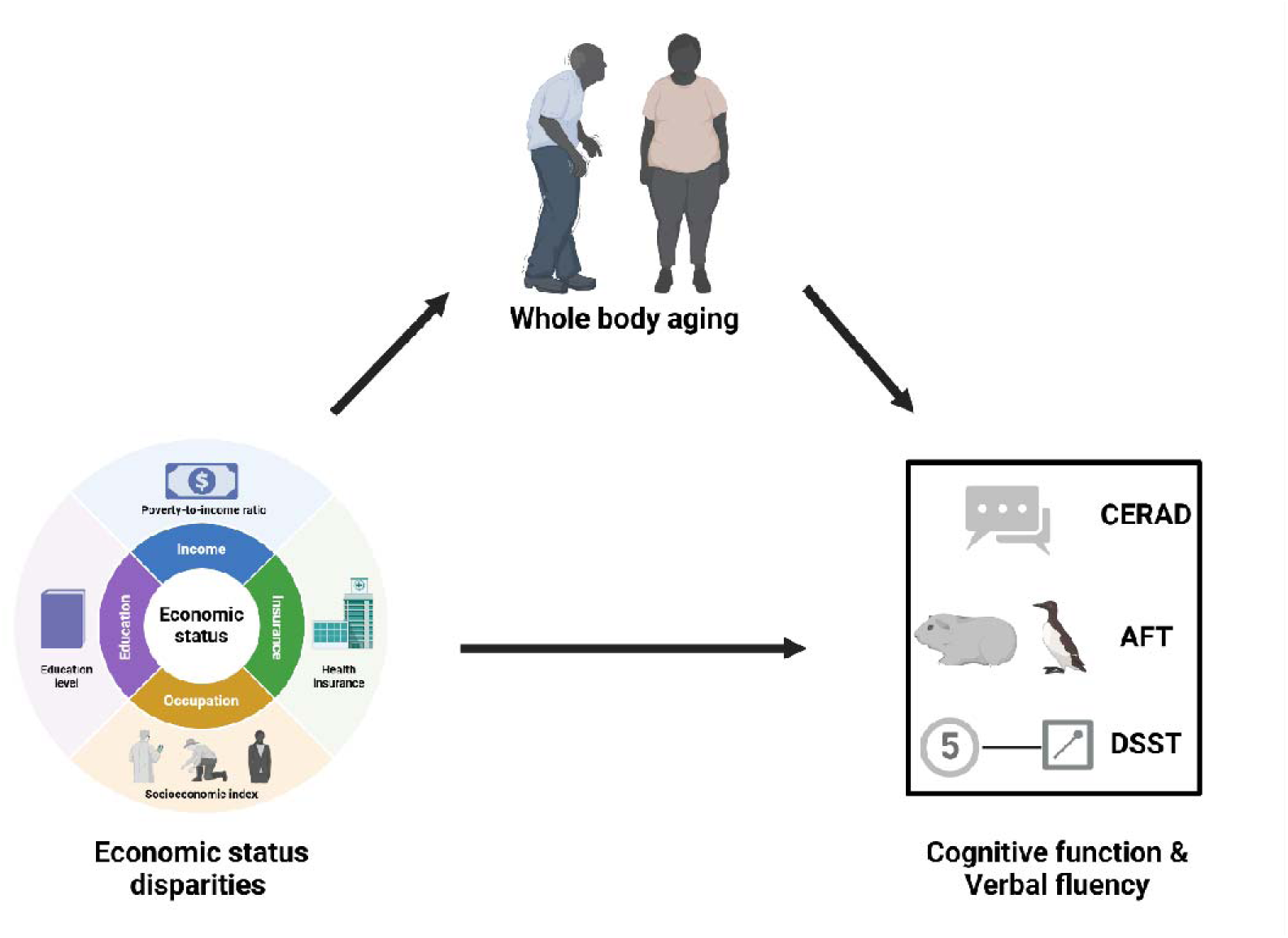

## 1. Introduction

Socioeconomic status (SES) disparities in income, education, and social status have been recognized as critical determinants in physical and cognitive functions, leading to changes in motor function, executive function, attention, learning, and verbal fluency, particularly in older adults (Boa Sorte Silva et al., 2024; Schwarz et al., 2024; Shan et al., 2024). Previous studies revealed that SES fosters brain structure and regulates cognitive functions by shaping gray and white matter volume and intrinsic connectivity (within the brain regions) (Cox et al., 2016; Montemurro et al., 2023; Morrison & Baxter, 2012). The deficits in cognitive performance have been acknowledged as a crucial concern among survivors of neurodegenerative disorders, such as Parkinsonism and Alzheimer’s disease and related dementias (Arenaza-Urquijo et al., 2024; Livingston et al., 2024). A meta-analysis covering 242,804 participants worldwide indicated that over 15% of community dwellers aged 50 years and older suffer from mild cognitive impairment (Bai et al., 2022). As populations age globally, understanding the intricate relationships between SES and cognitive decline becomes increasingly important (Christensen et al., 2009). The aging of the population structure increased vulnerability to cognitive impairment in memory, learning capacity, and verbal fluency, which are essential for role functioning and quality of life (Christensen et al., 2009).

Aging is an intricate process involving multiple biological changes, including telomere length shortening, epigenetic modifications, oxidative stress exacerbation, mitochondrial dysfunction, and cellular senescence (Santoro et al., 2021; Shim et al., 2024). Chronological age (CA) is generally considered an indicator of individual aging, however, it cannot manifest accurate physiological status and disease vulnerability. Thus, whole-body aging (WBA), which is based on multiple clinical biomarkers, provides a novel and holistic perspective for the recognition and intervention of aging-related disease. Researchers have also developed several clinical measures of WBA, including the Klemera-Doubal Age (Klemera & Doubal, 2006), Phenotypic Age (Liu et al., 2018), and Homeostatic Dysregulation (Cohen et al., 2013). Although WBA has been applied in predicting the incident and mortality risk of several age-related diseases, such as osteoarthritis, hypertensive diseases, diabetes, and cancer (Chen et al., 2022; Tian et al., 2023; Williams et al., 2023), the associations between WBA and cognitive function and verbal fluency remain unclear.

Despite great progress in the socioeconomic conditions and living standards in many countries and regions over recent decades, the gap in wealth inequity between developed and developing countries has become more pronounced. The inequities in SES contribute to disparities in health and survival—longevity, cognitive performance, and quality of life in middle- and high-income populations are better than those in low-income populations (Bor et al., 2017; “Mapping geographical inequalities in oral rehydration therapy coverage in low-income and middle-income countries, 2000-17,” 2020). Therefore, more efforts are warranted to reduce socioeconomic inequities in health and to improve the cognitive functions and quality of life in older populations.

Using data from the National Health and Nutrition Examination Survey (NHANES) 2011-2014, this study aims to explore the associations between SES disparities and cognitive functions and verbal fluency and the mediated effect of WBA among older adults, providing insights into the broader implications of SES-related cognitive declines in aging populations.

## 2. Methods

### 2.1. Study design and participants

NHANES is a nationally representative survey of the civilian, community-dwelling members in the United States. The survey was conducted by the Centers for Disease Control and Prevention (CDC) and the National Center for Health Statistics (NCHS) using a multistage probability design. Further details on study design have been reported previously (*Centers for Disease Control and Prevention/National Center for Health Statistics. About the National Health and Nutrition Examination Survey.* , 2017). Of the 19,931 participants, 9554, 1802, and 6251 were excluded due to SES data, WBA data, and/or cognitive function data missing, respectively. Finally, 2321 participants enrolled in the study (**Figure 1**). The NHANES was approved by the NCHS Research Ethics Review Board. All the participants provided written informed consent according to the research protocol.

**Figure 1.**
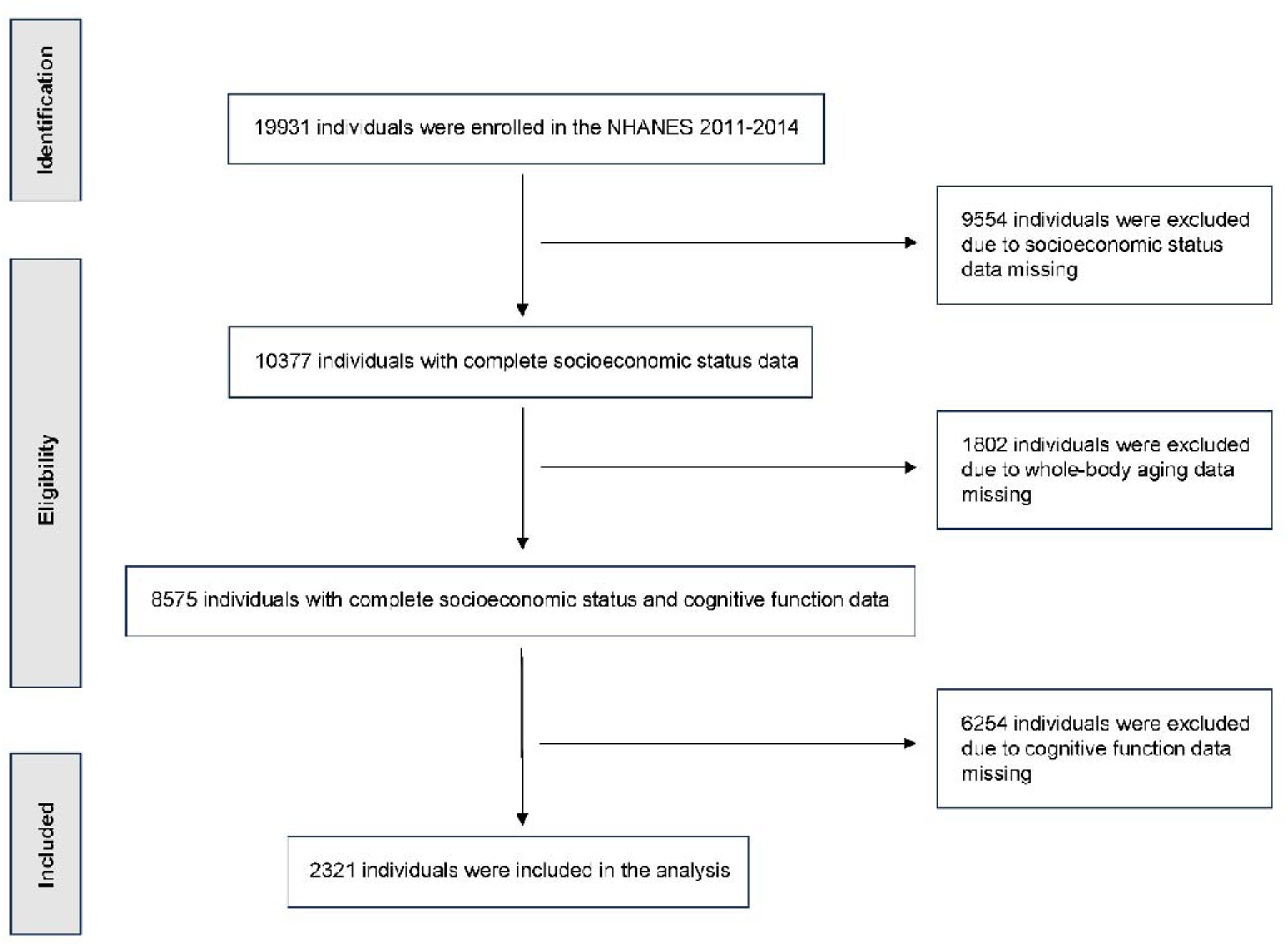
Enrollment process of this study.

### 2.2. Measurements

#### 2.2.1. Demographic data

The participants’ demographic data included their age (years), gender (male or female), ethnicity (Mexican American, other Hispanic, non-Hispanic White, non-Hispanic Black, non-Hispanic Asian, or other race), marital status (married, widowed/divorced/separated, never married, or living with a partner), body mass index (BMI) (< 18.5, 18.5-24.9, 25.0-29.9, ≥ 30.0), drinking (never, ever), smoking (never, ever), vigorous-intensity physical activity (yes, no).

#### 2.2.2. Socioeconomic status

The participants’ SES was evaluated by self-reported family income level, education, occupation, and health insurance according to previous studies (Quaglia et al., 2013; Zhang et al., 2021). These indicators were classified into three levels (low, moderate, and high). The family poverty-to-income ratio (PIR), which indicates the ratio of family annual income to the federal poverty level, was used to evaluate the level of family income. The score *S* of PIR was categorized as follows: *S* ≤ 1 = low family income level, 1 < *S*□< 4 = moderate family income level, and *S* ≥ 4 = high family income level. Education level was classified into three levels as well: less than senior high school diploma, senior high school graduate/GED, and equivalent, or college/university and above. The socioeconomic index based on the employee’s salary, education, and social prestige was used to evaluate the level of each occupation (Stevens & Cho, 1985). The levels of occupation were also classified into three categories: upper (socioeconomic index ≥ 50), lower (socioeconomic index < 50, including retirees and students), and unemployment (Zhang et al., 2021). Health insurance was classified into three categories: private insurance (including any private health insurance, Medi-Gap, or single-service plans), public insurance only (including Medicare, Medicaid, State Children’s Health Insurance Program, military healthcare, Indian Health Service, state-sponsored health plans, or other government programs), and uninsured (Le et al., 2020).

#### 2.2.3. Whole-body aging

WBA was calculated using three composite measures based on blood chemistry and clinical data: the Klemera-Doubal Method biological age (KDM) (Klemera & Doubal, 2006), PhenoAge (Levine et al., 2018), and homeostatic dysregulation (HD) (Cohen et al., 2013). The details of these measures for WBA have been previously reported otherwise (Graf et al., 2022; Kwon & Belsky, 2021). Briefly, KDM is derived from regressions of biomarkers on CA and reflects the age at which an individual’s physiology matches the average physiology of participants in the NHANES III. PhenoAge is based on a mortality prediction score using biomarkers and CA, representing the age at which an individual’s mortality risk matches the average risk in NHANES III. Unlike KDM and PhenoAge, HD does not incorporate CA in its calculation. Instead, it uses the Mahalanobis distance to quantify the deviation of a person’s physiology from a healthy population of NHANES III participants aged from 20 to 30.

Any age-related biomarkers can be used to construct biological age algorithms generally. We selected the same set of biomarkers for our KDM, PhenoAge, and HD algorithms to ensure comparability using the R package BioAge (Kwon & Belsky, 2021). We first identified 16 potential biomarkers, covering a range of organ systems (e.g., cardiometabolic, inflammatory, and kidney functions), that are routinely collected in clinical practice and available in NHANES III. We considered only biomarkers with ≤ 20% missing data and a correlation with chronological age (|r|□>□0.1, in line with prior research). The 16 age-related biomarkers and distribution for constructing KDM, PhenoAge, and HD algorithms are shown in **Figure S1**. Following previous studies, we selected non-pregnant participants aged 30–75 years with complete biomarker data as the reference population for KDM (n□=□7,694) (Mak et al., 2023). The reference population for PhenoAge included participants aged 20–84 years with complete biomarker data (n□=□12,998), while for HD, we selected participants aged 20–30 years who were not obese and had biomarker values within the age- and sex-specific normal range (n□=□258). Each individual had only one measurement occasion in the training set. The newly trained algorithms were then projected onto the NHANES 2011-2014 dataset. The distributions and correlations of the WBA and CA in the present study are shown in **Figures S3 and S4**.

#### 2.2.4. Cognitive functions and verbal fluency

Immediate and delayed learning ability and inhibition for novel verbal information were assessed using the Consortium to Establish a Registry for Alzheimer’s Disease (CERAD) Word List Learning Test (WLLT), Word List Recall Test (WLRT), and Intrusion Word Count Test (WLLT-IC and WLRT-IC). Verbal fluency and executive function were assessed using the Animal Fluency Test (AFT), and sustained attention, processing speed, and working memory were assessed using the Digit Symbol Substitution Test (DSST). Scoring in the CERAD WLLT, WLLT-IC, WLRT, and WLRT-IC ranged between 0 and 10, in AFT from 1 to 40, and in DSST from 0 to 100. Lower scores on cognitive tests above indicate more severe cognitive impairment. These tests have been previously validated for use in research practice.

### 2.3. Statistical analysis

SAS procedure PROC LCA was used to generate an unmeasured variable (SES) from multiple observed exclusive categorical variables (Lanza et al., 2007). An overall SES variable was calculated using latent class analysis based on four individual socioeconomic factors (family income level, education, occupation, and health insurance) (Quaglia et al., 2013). The SES levels of participants were divided into three latent classes (high, moderate, and low SES) according to the posterior probabilities. The details of the latent class analysis can be found in the **Table S1** and **Figure S2**.

Descriptive statistical analyses were conducted to exhibit the participants’ demographic profile and SES levels. Independent t-tests, one-way ANOVA tests, and Spearman correlation analysis were used to examine the associations between demographic variables, SES, whole-body aging, and cognitive functions. Multiple regression analysis was used to investigate the adjusted associations between SES, whole-body aging, and cognitive functions. The potential mediating effects of whole-body aging on the associations of SES and cognitive functions were estimated by simple mediation models using the bootstrap method with 1000 simulations (Montoya & Hayes, 2017; Valente et al., 2020). The total effect (TE) indicated the overall effect of SES on cognitive functions. The direct effect (DE) indicated the effects of SES on cognitive functions without whole-body aging. The indirect effect (IE) indicated the effects of SES on cognitive functions through whole-body aging. The mediation proportion = IE/TE.

The analyses were performed using SAS version 9.4 (SAS Institute, Cary, NC) and R 4.1.3. Two-sided *p* values < 0.05 were considered to be significant.

## 3. Results

### 3.1. Demographic characteristics

Among 2321 participants (mean age 69.3 years, 50.5% female), 561 (24.2%) were of low SES, 1142 (49.4%) were of moderate SES, and 618 (26.5%) were of high SES. The ethnicity of most participants was non-Hispanic White (1151, 49.6%), followed by non-Hispanic Black (527, 22.7%). Most participants were married or cohabiting (889, 57.4%). The demographic characteristics of BMI, drinking, smoking, and physical activity are listed in **Table 1**.

**Table 1.** Socio-demographic characteristics of participants (*n*= 2321).

| Variables | *n* | Percentage (%) |
| --- | --- | --- |
| **Age** |  |  |
| 60-69 years | 1284 | 55.3 |
| 70-79 years | 681 | 29.3 |
| ≥ 80 years | 356 | 15.4 |
| **Gender** |  |  |
| Male | 1149 | 49.5 |
| Female | 1172 | 50.5 |
| **Ethnicity** |  |  |
| Mexican American | 197 | 8.5 |
| Other Hispanic | 239 | 10.3 |
| Non-Hispanic White | 1151 | 49.6 |
| Non-Hispanic Black | 527 | 22.7 |
| Non-Hispanic Asian | 177 | 7.6 |
| Other Race - Including Multi-Racial | 30 | 1.3 |
| **Marital status <sup>a</sup>** |  |  |
| Married or cohabiting | 1364 | 58.7 |
| Widowed | 425 | 18.3 |
| Divorced | 322 | 14.3 |
| Separated | 66 | 2.8 |
| Never married | 133 | 5.7 |
| **Body mass index, kg/m<sup>2</sup> <sup>b</sup>** |  |  |
| < 18.5 | 30 | 1.3 |
| 18.5-24.9 | 578 | 24.9 |
| 25.0-29.9 | 825 | 35.5 |
| ≥ 30 | 875 | 37.7 |
| **Drinking <sup>c</sup>** |  |  |
| Never | 336 | 14.5 |
| Ever | 1959 | 84.4 |
| **Smoking <sup>d</sup>** |  |  |
| Never | 1130 | 48.7 |
| Ever | 1189 | 51.2 |
| **Vigorous-intensity physical activity <sup>e</sup>** |  |  |
| Yes | 271 | 11.7 |
| No | 2049 | 88.3 |
| **Socioeconomic status** |  |  |
| Low (Class 1) | 561 | 24.2 |
| Moderate (Class 2) | 1142 | 49.4 |
| High (Class 3) | 618 | 26.5 |

<sup>a</sup> 1 participant refused to respond; <sup>b</sup> Data was missing from 13 participants; <sup>c</sup> 5 participants refused to respond and data was missing from 21 participants; <sup>d</sup> 2 participants refused to respond; <sup>e</sup> 1 participants refused to respond.

### 3.2. Associations between demographic characteristics, socioeconomic status, and cognitive functions

**Table 2** shows the univariate analysis of associations between demographic characteristics and socioeconomic status with cognitive functions. Age, ethnicity, marital status, and SES were associated with CERAD WLLT (all *p* < 0.01), CERAD WLLRT (all *p* < 0.01), CERAD WLLT-IC (all *p* < 0.01), CERAD WLLRT-IC (all *p* < 0.01), AFT (all *p* < 0.01), and DSST (all *p* < 0.01). Gender was associated with CERAD WLLT (all *p* < 0.01), CERAD WLLRT (all *p* < 0.01), CERAD WLLT-IC (all *p* < 0.01), CERAD WLLRT-IC (all *p* < 0.01) and DSST (all *p* < 0.01) except for AFT (*t* =1.63, *p* = 0.102).

**Table 2.** Univariate analysis of the associations between demographic characteristics and socioeconomic status with cognitive functions.

|  | CERAD WLLT |  | CERAD WLLRT |  | CERAD WLLT-IC |  | CERAD WLLRT-IC |  | AFT |  | DSST |  |
| --- | --- | --- | --- | --- | --- | --- | --- | --- | --- | --- | --- | --- |
|  | *F / t* | *p* | *F / t* | *p* | *F / t* | *p* | *F / t* | *p* | *F / t* | *p* | *F / t* | *p* |
| Age | 50.86 | <0.001 | 56.12 | <0.001 | 46.11 | <0.001 | 76.78 | <0.001 | 39.22 | <0.001 | 71.06 | <0.001 |
| Gender | 6.94 | <0.001 | 8.34 | <0.001 | 7.21 | <0.001 | 8.11 | <0.001 | 1.63 | 0.102 | 7.07 | <0.001 |
| Ethnicity | 7.57 | <0.001 | 4.82 | <0.001 | 2.77 | 0.017 | 8.84 | <0.001 | 31.99 | <0.001 | 58.24 | <0.001 |
| Marital status | 4.56 | <0.001 | 4.47 | <0.001 | 3.20 | 0.004 | 3.73 | 0.001 | 9.20 | <0.001 | 14.80 | <0.001 |
| Socioeconomic status | 50.86 | <0.001 | 56.12 | <0.001 | 46.11 | <0.001 | 76.78 | <0.001 | 39.22 | <0.001 | 71.06 | <0.001 |

Note: AFT, Animal Fluency Test; CERAD, Consortium to Establish a Registry for Alzheimer's Disease; DSST, Digit Symbol Substitution Test; WLLT, Word List Learning Test; WLLRT, Word List Recall Test.; WLLT-IC, Word List Learning
Test—Intrusion Word Count; WLRT-IC, Word List Recall Test—Intrusion Word Count. A two-tailed $p < 0.05$ was indicative of statistical significance.

**Table 3** shows the correlation analysis of the associations between WBA and cognitive functions. There were statistically significant positive correlations between KDM, PhenoAge, and HD. The WBA was negatively associated with cognitive functions and verbal fluency. The levels of SES were positively associated with CERAD WLLT (β *=*0.20, *P* <0.001), CERAD WLLRT (β *=*0.19, *P* <0.001), CERAD WLLT-IC (β *=*0.16, *P* <0.001), WLLRT-IC (β *=*0.17, *P* <0.001), AFT (β *=*0.28, *P* <0.001), and DSST (β *=*0.45, *P* <0.001) in the adjusted linear regression models. Moreover, the WBA was negatively associated with cognitive function and verbal fluency in the multivariate analysis (**Table 4**).

**Table 3.** Correlation analysis of the associations between whole-body aging and cognitive.

|  | 1 | 2 | 3 | 4 | 5 | 6 | 7 | 8 | 9 |
| --- | --- | --- | --- | --- | --- | --- | --- | --- | --- |
| 1. KDM | 1 |  |  |  |  |  |  |  |  |
| 2. PhenoAge | **0.70\*\*** | 1 |  |  |  |  |  |  |  |
| 3. HD | **0.57\*\*** | **0.36\*\*** | 1 |  |  |  |  |  |  |
| 4. CERAD WLLT | **-0.15\*\*** | **-0.21\*\*** | **-0.05\*** | 1 |  |  |  |  |  |
| 5. CERAD WLLRT | **-0.19\*\*** | **-0.25\*\*** | **-0.05\*** | **0.63\*\*** | 1 |  |  |  |  |
| 6. CERAD WLLT-IC | **-0.19\*\*** | **-0.24\*\*** | **-0.05\*** | **0.56\*\*** | **0.70\*\*** | 1 |  |  |  |
| 7. CERAD WLLRT-IC | **-0.21\*\*** | **-0.27\*\*** | **-0.05\*** | **0.59\*\*** | **0.67\*\*** | **0.70\*\*** | 1 |  |  |
| 8. AFT | **-0.15\*\*** | **-0.22\*\*** | **-0.14\*\*** | **0.31\*\*** | **0.36\*\*** | **0.35\*\*** | **0.34\*\*** | 1 |  |
| 9. DSST | **-0.27\*\*** | **-0.34\*\*** | **-0.13\*\*** | **0.42\*\*** | **0.44\*\*** | **0.40\*\*** | **0.44\*\*** | **0.51\*\*** | 1 |

Note: AFT, Animal Fluency Test; CERAD, Consortium to Establish a Registry for Alzheimer's Disease; DSST, Digit Symbol Substitution Test; Klemara-Doubal Method, KDM; Homeostatic dysregulation, HD; WLLT, Word List Learning Test; WLRT, Word List Recall Test.; WLLT-IC, Word List Learning Test—Intrusion Word Count; WLRT-IC, Word List Recall Test—Intrusion Word Count. \* $p < 0.05$ , \*\* $p < 0.01$ .

**Table 4.**
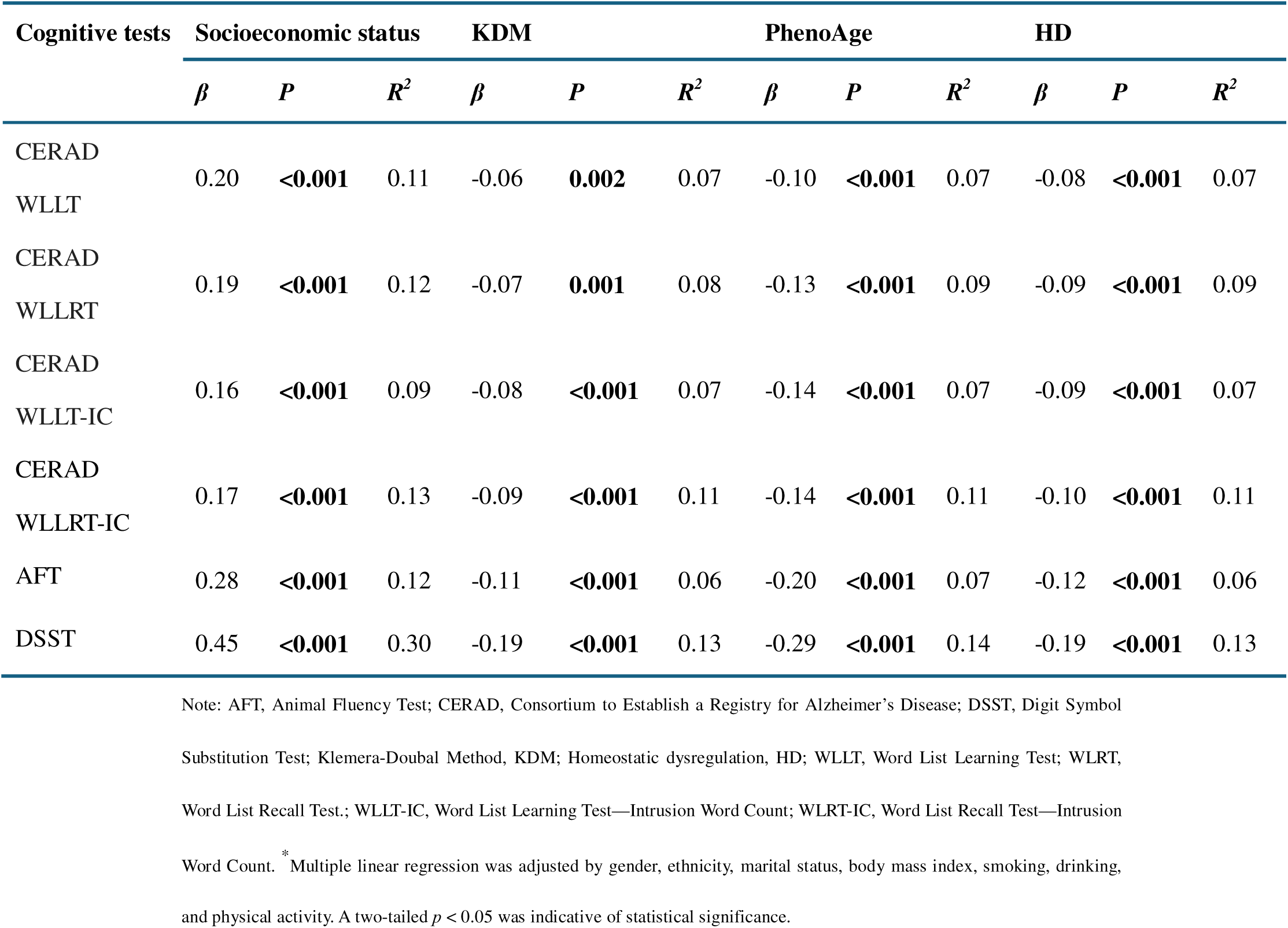
Multivariate analysis of the associations between socioeconomic status and whole-body aging with cognitive function.

### 3.3. Whole-body aging mediates the association between socioeconomic status and cognition and verbal fluency

**Figure 2** describes the mediation effects of WBA on the association between SES and cognitive functions and verbal fluency. After adjusting gender, ethnicity, marital status, body mass index, smoking, drinking, and physical activity in the mediation analysis, the proportions of indirect effects of SES and cognition and verbal fluency were 3.21%-5.17% for KDM. The proportions of indirect effects of SES and cognition and verbal fluency were 3.42%-7.65% for PhenoAge. The proportions of indirect effects of SES and cognition and verbal fluency were 3.77%-7.40% for HD. Moreover, no mediation effect of KDM on SES and CERAD WLLT was observed.

**Figure 2.**
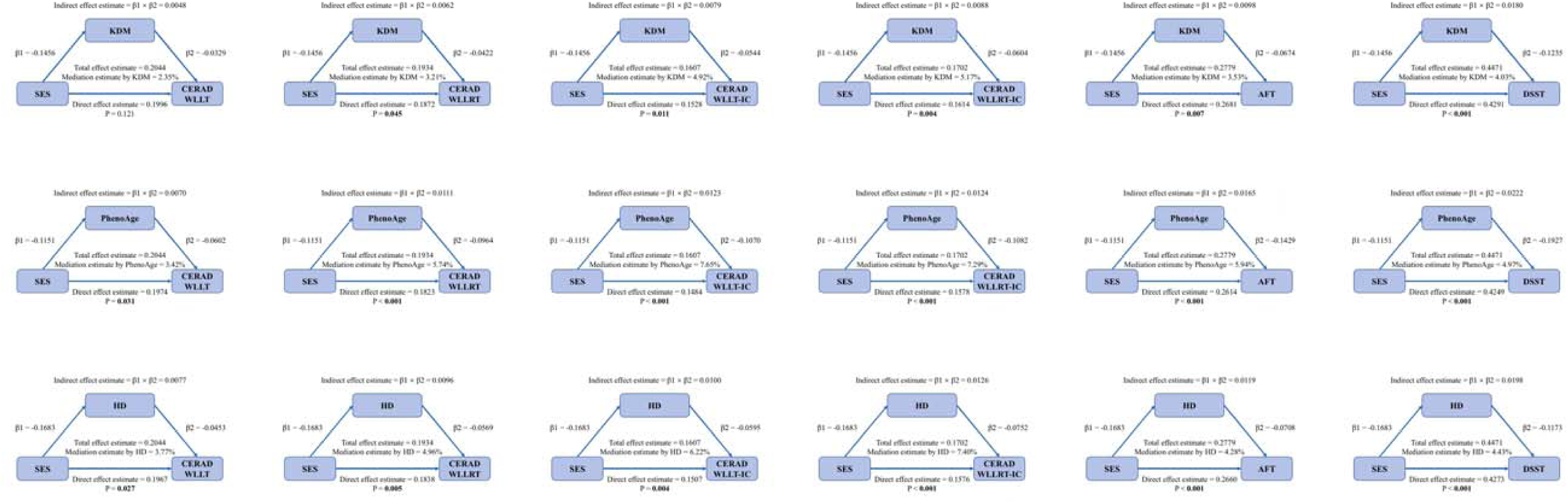
Whole-body aging mediates the association between socioeconomic status, cognitive functions, and verbal fluency. AFT, Animal Fluency Test; CERAD, Consortium to Establish a Registry for Alzheimer’s Disease; DSST, Digit Symbol Substitution Test; Homeostatic dysregulation, HD; Klemera-Doubal Method, KDM; Socioeconomic status, SES; WLLT, Word List Learning Test; WLRT, Word List Recall Test.; WLLT-IC, Word List g Test—Intrusion Word Count; WLRT-IC, Word List Recall Learning Test—Intrusion Word Count. Mediation analysis was adjusted by gender, ethnicity, marital status, body mass index, smoking, drinking, and physical activity. A two-tailed p < 0.05 was indicative of statistical significance.

## 4. Discussion

In the present study, we explore the relationship between SES and cognitive functions as well as verbal fluency among U.S. middle-to-old-aged adults. Our study demonstrated that higher SES was significantly associated with better cognitive functions and verbal fluency and accelerated WBA partially mediated the associations between SES and cognitive functions and verbal fluency. The findings reveal a novel pathway linking SES to cognitive decline and the significance of eliminating the SES disparities.

### Comparison with Previous Studies

Our findings are consistent with prior research (Wang et al., 2023), which highlights that SES strongly influences cognitive abilities, including immediate and delayed learning ability, sustained attention, processing speed, working memory, and verbal fluency. SES disparities encompass variations in income, education, occupation, and health insurance, which in turn shape access to resources such as healthcare utilization, nutritious food, and cognitively stimulating environments. Formal education is correlated with higher individual cognitive functions across the lifespan, and prolonging education attenuates aging-related cognitive declines (Du et al., 2023). Improving the educational conditions that shape development during the first decades of life has emerged as a great potential approach for preserving cognitive ability in old age and for reducing public burdens related to cognition-related diseases (Lövdén et al., 2020).

In terms of income, a cross-nationally harmonized longitudinal study indicated that wealth loss of 75% or greater in later life is negatively associated with subsequent cognitive functions in the USA and China (Cho et al., 2023). Similarly, economic downturns around retirement increase cognitive decline in later life, with the long-lasting effect persisting for up to 10 years (Hessel et al., 2018). Individuals with higher socioeconomic status perform slower cognitive decline among older Chinese immigrants in Chicago, which underscores the essential role of economic and occupational factors in cognitive functions (Tang et al., 2023). Moreover, health insurance also plays a vital role in aging-related cognitive decline. Studies from China, the USA, and European countries demonstrate that health insurance coverage is associated with a reduced risk of cognitive decline (Li et al., 2023; Peng et al., 2023; Wang et al., 2023). These findings indicated government policies and social safety nets should protect individuals from wealth losses in later life to reduce cognitive decline and related diseases.

Among the 18 mediation analysis models, 17 exhibited a significant mediated effect of WBA between SES and cognitive declines. The findings revealed that WBA accounts for a modest but statistically significant portion of the association between SES and cognitive ability. SES makes an impact on cognition both directly, through psychosocial mechanisms, and indirectly, by accelerating physiological aging. The mediation effect emphasizes the importance of addressing physiological aging in interventions aimed at mitigating SES-related cognitive disparities. Targeting modifiable risk factors such as diet, physical activity, and stress management could potentially slow WBA and preserve cognitive ability.

### Implications for Policy and Practice

The findings have critical implications for public health policies and interventions aimed at reducing health disparities. Policymakers should prioritize strategies to address SES-related inequities, such as expanding access to income, high-quality education, and healthcare utilization for socioeconomically disadvantaged groups. Incorporating WBA evaluation into routine clinical assessments could enable early identification of individuals at risk for accelerated aging and cognitive decline. Furthermore, community-based programs promoting lifestyle modifications—such as physical activity, stress management, and healthy diets—can help mitigate WBA and preserve cognitive function. These efforts should also include targeted initiatives for low-SES populations, addressing both the social determinants of health and the biological mechanisms contributing to disparities.

### Strengths and Limitations

To the best of our knowledge, few studies have examined the mediating role of WBA in the relationship between SES and cognitive functions and verbal fluency. In this study, composite SES ranks and WBA status were constructed using multiple validated and robust measures, including the Klemera-Doubal Method biological aging, phenotypic aging, and homeostatic dysregulation. We investigated both direct and indirect associations between SES disparities, WBA, cognitive function, and verbal fluency in a representative U.S. population. Additionally, the inclusion of multiple cognitive domains allowed for a comprehensive analysis of SES-related disparities in cognitive functions and verbal fluency. These efforts enhance the generalizability and robustness of our findings.

However, certain limitations also should be considered in the study. First, the cross-sectional nature of the study design precludes causal inference, limiting our ability to establish temporal relationships between SES, WBA, and cognitive functions. Longitudinal studies are needed to verify the observed associations and explore potential bidirectional effects. Second, self-reported SES measures may introduce reporting bias. Third, although we used a representative U.S. cohort in the study, external validation in other cohorts is warranted given the different developmental status and health-related characteristics across countries and regions. Finally, while our study used well-validated WBA measures, other biological markers, such as epigenetic modification or telomere length, may further elucidate the SES-Aging-Cognition pathway.

## 5. Conclusion

In conclusion, the study suggested that SES disparities were significantly associated with immediate and delayed learning ability, sustained attention, processing speed, working memory, and verbal fluency, which was possibly and partially mediated by WBA. By elucidating the interplay between SES and WBA, the findings contribute to a deeper understanding of cognitive decline and underscore the importance of addressing socioeconomic inequities and physiological aging processes to promote cognitive resilience among older populations. Future research should prioritize longitudinal investigations of these associations and explore further biological mechanisms to explore comprehensive interventions for cognitive health equity.

## Declaration of Interest Statement

The authors have no conflicts to disclose.

## Data Availability

NHANES is an open-access resource. All researchers can apply to use its data for health-related research that is in the public interest (https://wwwn.cdc.gov/nchs/nhanes/Default.aspx).

## Acknowledgments

None.

## Funding statement

The study was supported by the National Social Science Fund of China to Y.X. (Award number: 13CYY023).

## Human ethics and consent to participate declarations

The NCHS Research Ethics Review Board approved the NHANES in accordance with the Declaration of Helsinki. All the participants provided written informed consent according to the research protocol.

## Clinical trial number

The study is an observational study. Clinical trial registration number is not applicable.

## Consent for publication

Not applicable.

## Author Contributions

Y.X. drafted the structure of the present work, conducted data analysis, drafted the first version of the manuscript, and revised subsequent versions critically for important intellectual content. L.X. revised the manuscript. Y.X. and W.S. designed the study. All authors had full access to all data in the study, revised the work critically for important intellectual content, gave final approval of the version to be published, and agreed to be accountable for all aspects of the work in ensuring that questions related to the accuracy or integrity of any part were appropriately investigated and resolved.

## Notes

### Competing Interest Statement

The authors have declared no competing interest.

## Reference

Arenaza-Urquijo, E. M., Boyle, R., Casaletto, K., Anstey, K. J., Vila-Castelar, C., Colverson, A., Palpatzis, E., Eissman, J. M., Kheng Siang Ng, T., Raghavan, S., Akinci, M., Vonk, J. M. J., Machado, L. S., Zanwar, P. P., Shrestha, H. L., Wagner, M., Tamburin, S., Sohrabi, H. R., Loi, S.,…Buckley, R. F. (2024). Sex and gender differences in cognitive resilience to aging and Alzheimer’s disease. Alzheimers Dement. 10.1002/alz.13844

Bai, W., Chen, P., Cai, H., Zhang, Q., Su, Z., Cheung, T., Jackson, T., Sha, S., & Xiang, Y. T. (2022). Worldwide prevalence of mild cognitive impairment among community dwellers aged 50 years and older: a meta-analysis and systematic review of epidemiology studies. Age Ageing, 51(8). 10.1093/ageing/afac173

Boa Sorte Silva, N. C., Barha, C. K., Erickson, K. I., Kramer, A. F., & Liu-Ambrose, T. (2024). Physical exercise, cognition, and brain health in aging. Trends Neurosci, 47(6), 402–417. 10.1016/j.tins.2024.04.004

Bor, J., Cohen, G. H., & Galea, S. (2017). Population health in an era of rising income inequality: USA, 1980-2015. Lancet, 389(10077), 1475–1490. 10.1016/s0140-6736(17)30571-8

*Centers for Disease Control and Prevention/National Center for Health Statistics. About the National Health and Nutrition Examination Survey.*. (2017). www.cdc.gov/nchs/nhanes/about_nhanes.htm

Chen, L., Zhao, Y., Liu, F., Chen, H., Tan, T., Yao, P., & Tang, Y. (2022). Biological aging mediates the associations between urinary metals and osteoarthritis among U.S. adults. BMC Med, 20(1), 207. 10.1186/s12916-022-02403-3

Cho, T. C., Yu, X., Gross, A. L., Zhang, Y. S., Lee, J., Langa, K. M., & Kobayashi, L. C. (2023). Negative wealth shocks in later life and subsequent cognitive function in older adults in China, England, Mexico, and the USA, 2012-18: a population-based, cross-nationally harmonised, longitudinal study. Lancet Healthy Longev, 4(9), e461–e469. 10.1016/s2666-7568(23)00113-7

Christensen, K., Doblhammer, G., Rau, R., & Vaupel, J. W. (2009). Ageing populations: the challenges ahead. Lancet, 374(9696), 1196–1208. 10.1016/s0140-6736(09)61460-4

Cohen, A. A., Milot, E., Yong, J., Seplaki, C. L., Fülöp, T., Bandeen-Roche, K., & Fried, L. P. (2013). A novel statistical approach shows evidence for multi-system physiological dysregulation during aging. Mech Ageing Dev, 134(3-4), 110–117. 10.1016/j.mad.2013.01.004

Cox, S. R., Dickie, D. A., Ritchie, S. J., Karama, S., Pattie, A., Royle, N. A., Corley, J., Aribisala, B. S., Valdés Hernández, M., Muñoz Maniega, S., Starr, J. M., Bastin, M. E., Evans, A. C., Wardlaw, J. M., & Deary, I. J. (2016). Associations between education and brain structure at age 73 years, adjusted for age 11 IQ. Neurology, 87(17), 1820–1826. 10.1212/wnl.0000000000003247

Du, C., Miyazaki, Y., Dong, X., & Li, M. (2023). Education, Social Engagement, and Cognitive Function: A Cross-Lagged Panel Analysis. J Gerontol B Psychol Sci Soc Sci, 78(10), 1756–1764. 10.1093/geronb/gbad088

Graf, G. H., Crowe, C. L., Kothari, M., Kwon, D., Manly, J. J., Turney, I. C., Valeri, L., & Belsky, D. W. (2022). Testing Black-White Disparities in Biological Aging Among Older Adults in the United States: Analysis of DNA-Methylation and Blood-Chemistry Methods. Am J Epidemiol, 191(4), 613–625. 10.1093/aje/kwab281

Hessel, P., Riumallo-Herl, C. J., Leist, A. K., Berkman, L. F., & Avendano, M. (2018). Economic Downturns, Retirement and Long-Term Cognitive Function Among Older Americans. J Gerontol B Psychol Sci Soc Sci, 73(4), 744–754. 10.1093/geronb/gbx035

Klemera, P., & Doubal, S. (2006). A new approach to the concept and computation of biological age. Mech Ageing Dev, 127(3), 240–248. 10.1016/j.mad.2005.10.004

Kwon, D., & Belsky, D. W. (2021). A toolkit for quantification of biological age from blood chemistry and organ function test data: BioAge. Geroscience, 43(6), 2795–2808. 10.1007/s11357-021-00480-5

Lanza, S. T., Collins, L. M., Lemmon, D. R., & Schafer, J. L. (2007). PROC LCA: A SAS Procedure for Latent Class Analysis. Struct Equ Modeling, 14(4), 671–694. 10.1080/10705510701575602

Le, P., Chaitoff, A., Misra-Hebert, A. D., Ye, W., Herman, W. H., & Rothberg, M. B. (2020). Use of Antihyperglycemic Medications in U.S. Adults: An Analysis of the National Health and Nutrition Examination Survey. Diabetes Care, 43(6), 1227–1233. 10.2337/dc19-2424

Levine, M. E., Lu, A. T., Quach, A., Chen, B. H., Assimes, T. L., Bandinelli, S., Hou, L., Baccarelli, A. A., Stewart, J. D., Li, Y., Whitsel, E. A., Wilson, J. G., Reiner, A. P., Aviv, A., Lohman, K., Liu, Y., Ferrucci, L., & Horvath, S. (2018). An epigenetic biomarker of aging for lifespan and healthspan. Aging (Albany NY), 10(4), 573–591. 10.18632/aging.101414

Li, C.-S., Lai, G. C., Tsendsuren, S., Butler, R. J., & Liu, C.-C. (2023). Cognitive abilities and life insurance holdings: evidence from 16 European countries. The Geneva Risk and Insurance Review, 48(1), 110–166. 10.1057/s10713-022-00077-8

Liu, Z., Kuo, P. L., Horvath, S., Crimmins, E., Ferrucci, L., & Levine, M. (2018). A new aging measure captures morbidity and mortality risk across diverse subpopulations from NHANES IV: A cohort study. PLoS Med, 15(12), e1002718. 10.1371/journal.pmed.1002718

Livingston, G., Huntley, J., Liu, K. Y., Costafreda, S. G., Selbæk, G., Alladi, S., Ames, D., Banerjee, S., Burns, A., Brayne, C., Fox, N. C., Ferri, C. P., Gitlin, L. N., Howard, R., Kales, H. C., Kivimäki, M., Larson, E. B., Nakasujja, N., Rockwood, K.,…Mukadam, N. (2024). Dementia prevention, intervention, and care: 2024 report of the Lancet standing Commission. Lancet, 404(10452), 572–628. 10.1016/s0140-6736(24)01296-0

Lövdén, M., Fratiglioni, L., Glymour, M. M., Lindenberger, U., & Tucker-Drob, E. M. (2020). Education and Cognitive Functioning Across the Life Span. Psychol Sci Public Interest, 21(1), 6–41. 10.1177/1529100620920576

Mak, J. K. L., McMurran, C. E., Kuja-Halkola, R., Hall, P., Czene, K., Jylhävä, J., & Hägg, S. (2023). Clinical biomarker-based biological aging and risk of cancer in the UK Biobank. Br J Cancer, 129(1), 94–103. 10.1038/s41416-023-02288-w Mapping geographical inequalities in oral rehydration therapy coverage in low-income and middle-income countries, 2000-17. (2020). *Lancet Glob Health*, *8*(8), e1038-e1060. 10.1016/s2214-109x(20)30230-8

Montemurro, S., Filippini, N., Ferrazzi, G., Mantini, D., Arcara, G., & Marino, M. (2023). Education differentiates cognitive performance and resting state fMRI connectivity in healthy aging. Front Aging Neurosci, 15, 1168576. 10.3389/fnagi.2023.1168576

Montoya, A. K., & Hayes, A. F. (2017). Two-condition within-participant statistical mediation analysis: A path-analytic framework. Psychol Methods, 22(1), 6–27. 10.1037/met0000086

Morrison, J. H., & Baxter, M. G. (2012). The ageing cortical synapse: hallmarks and implications for cognitive decline. Nat Rev Neurosci, 13(4), 240–250. 10.1038/nrn3200

Peng, C., Burr, J. A., & Han, S. H. (2023). Cognitive function and cognitive decline among older rural Chinese adults: the roles of social support, pension benefits, and medical insurance. Aging Ment Health, 27(4), 771–779. 10.1080/13607863.2022.2088693

Quaglia, A., Lillini, R., Mamo, C., Ivaldi, E., & Vercelli, M. (2013). Socio-economic inequalities: a review of methodological issues and the relationships with cancer survival. Crit Rev Oncol Hematol, 85(3), 266–277. 10.1016/j.critrevonc.2012.08.007

Santoro, A., Bientinesi, E., & Monti, D. (2021). Immunosenescence and inflammaging in the aging process: age-related diseases or longevity? Ageing Res Rev, 71, 101422. 10.1016/j.arr.2021.101422

Schwarz, C., Franz, C. E., Kremen, W. S., & Vuoksimaa, E. (2024). Reserve, resilience and maintenance of episodic memory and other cognitive functions in aging. Neurobiol Aging, 140, 60–69. 10.1016/j.neurobiolaging.2024.04.011

Shan, Q., Yu, X., Lin, X., & Tian, Y. (2024). Reduced inhibitory synaptic transmission onto striatopallidal neurons may underlie aging-related motor skill deficits. Neurobiol Dis, 199, 106582. 10.1016/j.nbd.2024.106582

Shim, H. S., Iaconelli, J., Shang, X., Li, J., Lan, Z. D., Jiang, S., Nutsch, K., Beyer, B. A., Lairson, L. L., Boutin, A. T., Bollong, M. J., Schultz, P. G., & DePinho, R. A. (2024). TERT activation targets DNA methylation and multiple aging hallmarks. Cell, 187(15), 4030–4042.e4013. 10.1016/j.cell.2024.05.048

Stevens, G., & Cho, J. H. (1985). Socioeconomic indexes and the new 1980 census occupational classification scheme. Social Science Research, 14(2), 142–168. 10.1016/0049-089X(85)90008-0

Tang, F., Li, K., Rosso, A. L., Jiang, Y., & Li, M. (2023). Neighborhood segregation, socioeconomic status, and cognitive function among older Chinese immigrants. J Am Geriatr Soc, 71(3), 916–926. 10.1111/jgs.18167

Tian, Y. E., Cropley, V., Maier, A. B., Lautenschlager, N. T., Breakspear, M., & Zalesky, A. (2023). Heterogeneous aging across multiple organ systems and prediction of chronic disease and mortality. Nat Med, 29(5), 1221–1231. 10.1038/s41591-023-02296-6

Valente, M. J., Rijnhart, J. J. M., Smyth, H. L., Muniz, F. B., & MacKinnon, D. P. (2020). Causal Mediation Programs in R, Mplus, SAS, SPSS, and Stata. Struct Equ Modeling, 27(6), 975–984. 10.1080/10705511.2020.1777133

Wang, X., Bakulski, K. M., Paulson, H. L., Albin, R. L., & Park, S. K. (2023). Associations of healthy lifestyle and socioeconomic status with cognitive function in U.S. older adults. Scientific Reports, 13(1), 7513. 10.1038/s41598-023-34648-0

Williams, A. M., Mandelblatt, J. S., Wang, M., Dong, Q., Armstrong, G. T., Bhakta, N., Brinkman, T. M., Ehrhardt, M. J., Mulrooney, D. A., Gilmore, N., Robison, L. L., Yasui, Y., Small, B. J., Srivastava, D., Hudson, M. M., Ness, K. K., Krull, K. R., & Wang, Z. (2023). Deficit Accumulation Index and Biological Markers of Aging in Survivors of Childhood Cancer. JAMA Netw Open, 6(11), e2344015. 10.1001/jamanetworkopen.2023.44015

Zhang, Y. B., Chen, C., Pan, X. F., Guo, J., Li, Y., Franco, O. H., Liu, G., & Pan, A. (2021). Associations of healthy lifestyle and socioeconomic status with mortality and incident cardiovascular disease: two prospective cohort studies. Bmj, 373, n604. 10.1136/bmj.n604

