## Supplemental materials for "Whole-body aging mediates the association between socioeconomic status disparities and cognition and verbal fluency among U.S. older adults"

**Table S1.** Class membership and item-response probabilities in models with two to four latent classes.

| **Item** | **Latent class 1** | **Latent class 2** | **Latent class 3** | **Latent class 4** |
| --- | --- | --- | --- | --- |
| **Two-latent-class solution** | | | | |
| Mean posterior probabilities | 0.86 | 0.89 |  |  |
| Class membership probabilities | 0.48 | 0.52 | NA | NA |
| Income 1 | 0.35 | 0.00 | NA | NA |
| Income 2 | **0.65** | 0.48 | NA | NA |
| Income 3 | 0.00 | **0.52** | NA | NA |
| Occupation 1 | 0.08 | 0.03 | NA | NA |
| Occupation 2 | **0.48** | **0.51** | NA | NA |
| Occupation 3 | 0.44 | 0.46 | NA | NA |
| Education 1 | **0.43** | 0.06 | NA | NA |
| Education 2 | 0.29 | 0.18 | NA | NA |
| Education 3 | 0.27 | **0.76** | NA | NA |
| Insurance 1 | 0.16 | 0.05 | NA | NA |
| Insurance 2 | **0.56** | 0.17 | NA | NA |
| Insurance 3 | 0.27 | **0.78** | NA | NA |
| **Three-latent-class solution** | | | | |
| Mean posterior probabilities | 0.85 | 0.81 | 0.86 |  |
| Class membership probabilities | 0.29 | 0.47 | 0.23 | NA |
| Income 1 | 0.48 | 0.05 | 0.00 | NA |
| Income 2 | **0.51** | **0.86** | 0.02 | NA |
| Income 3 | 0.01 | 0.09 | **0.98** | NA |
| Occupation 1 | 0.09 | 0.04 | 0.02 | NA |
| Occupation 2 | **0.51** | 0.45 | **0.56** | NA |
| Occupation 3 | 0.40 | **0.50** | 0.42 | NA |
| Education 1 | **0.55** | 0.14 | 0.05 | NA |
| Education 2 | 0.24 | 0.29 | 0.11 | NA |
| Education 3 | 0.21 | **0.56** | **0.84** | NA |
| Insurance 1 | 0.19 | 0.08 | 0.04 | NA |
| Insurance 2 | **0.65** | 0.28 | 0.14 | NA |
| Insurance 3 | 0.16 | **0.64** | **0.82** | NA |
| **Four-latent-class solution** | | | | |
| Mean posterior probabilities | 0.73 | 0.75 | 0.60 | 0.73 |
| Class membership probabilities | 0.25 | 0.27 | 0.17 | 0.31 |
| Income 1 | **0.51** | 0.12 | 0.02 | 0.00 |
| Income 2 | 0.49 | **0.82** | **0.80** | 0.26 |
| Income 3 | 0.00 | 0.06 | 0.18 | **0.74** |
| Occupation 1 | 0.10 | 0.00 | 0.12 | 0.02 |
| Occupation 2 | **0.55** | 0.17 | **0.87** | **0.52** |
| Occupation 3 | 0.35 | **0.82** | 0.00 | 0.45 |
| Education 1 | 0.18 | 0.16 | 0.21 | 0.03 |
| Education 2 | **0.66** | 0.30 | **0.40** | 0.14 |
| Education 3 | 0.15 | **0.54** | 0.39 | **0.83** |
| Insurance 1 | 0.20 | 0.03 | 0.04 | 0.15 |
| Insurance 2 | **0.67** | **0.62** | **0.80** | 0.19 |
| Insurance 3 | 0.13 | 0.35 | 0.16 | **0.65** |

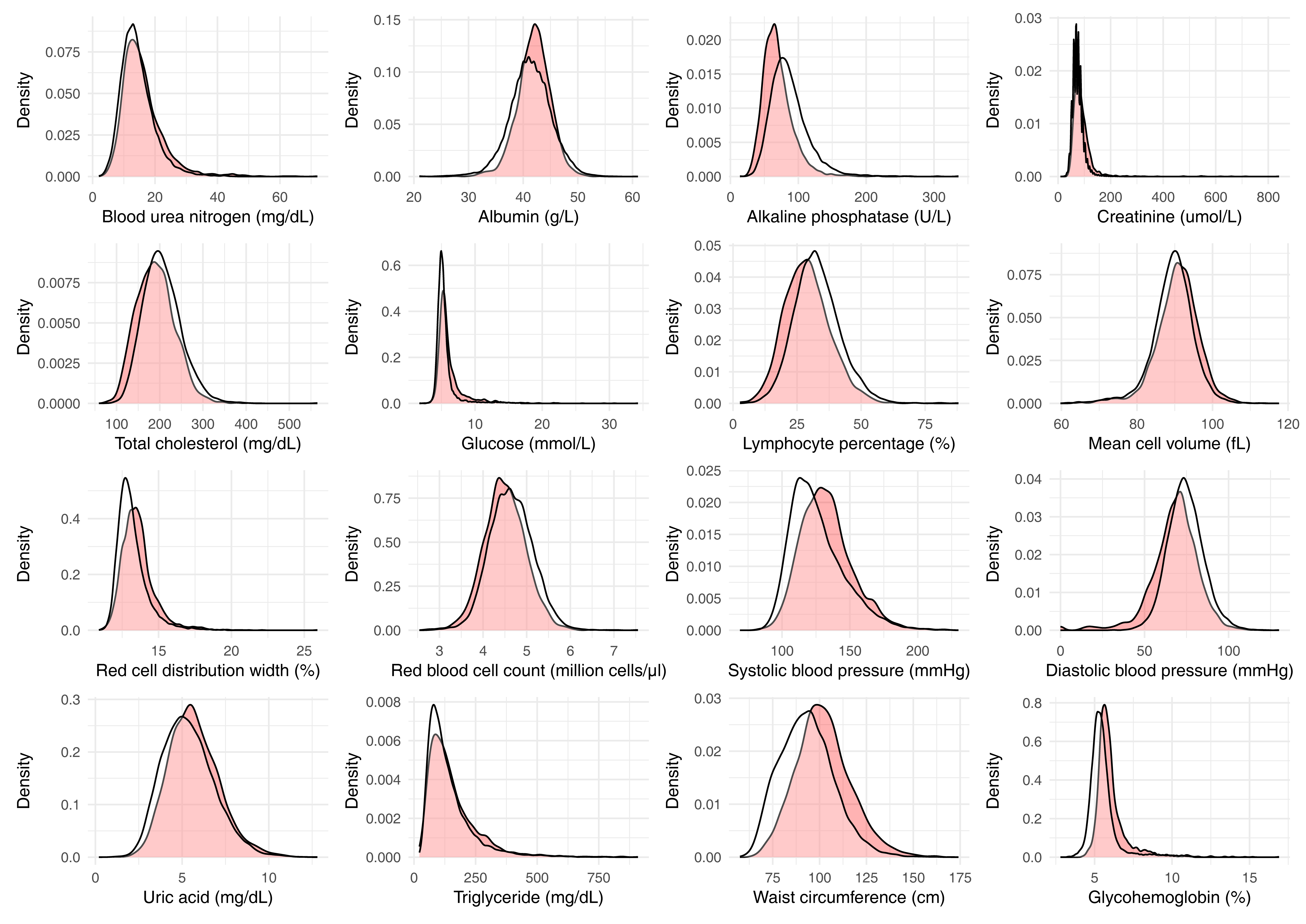

**Figure S1.** Distribution of biomarkers in individuals in NHANES III (white) and NHANES 2011-2014 (pink).

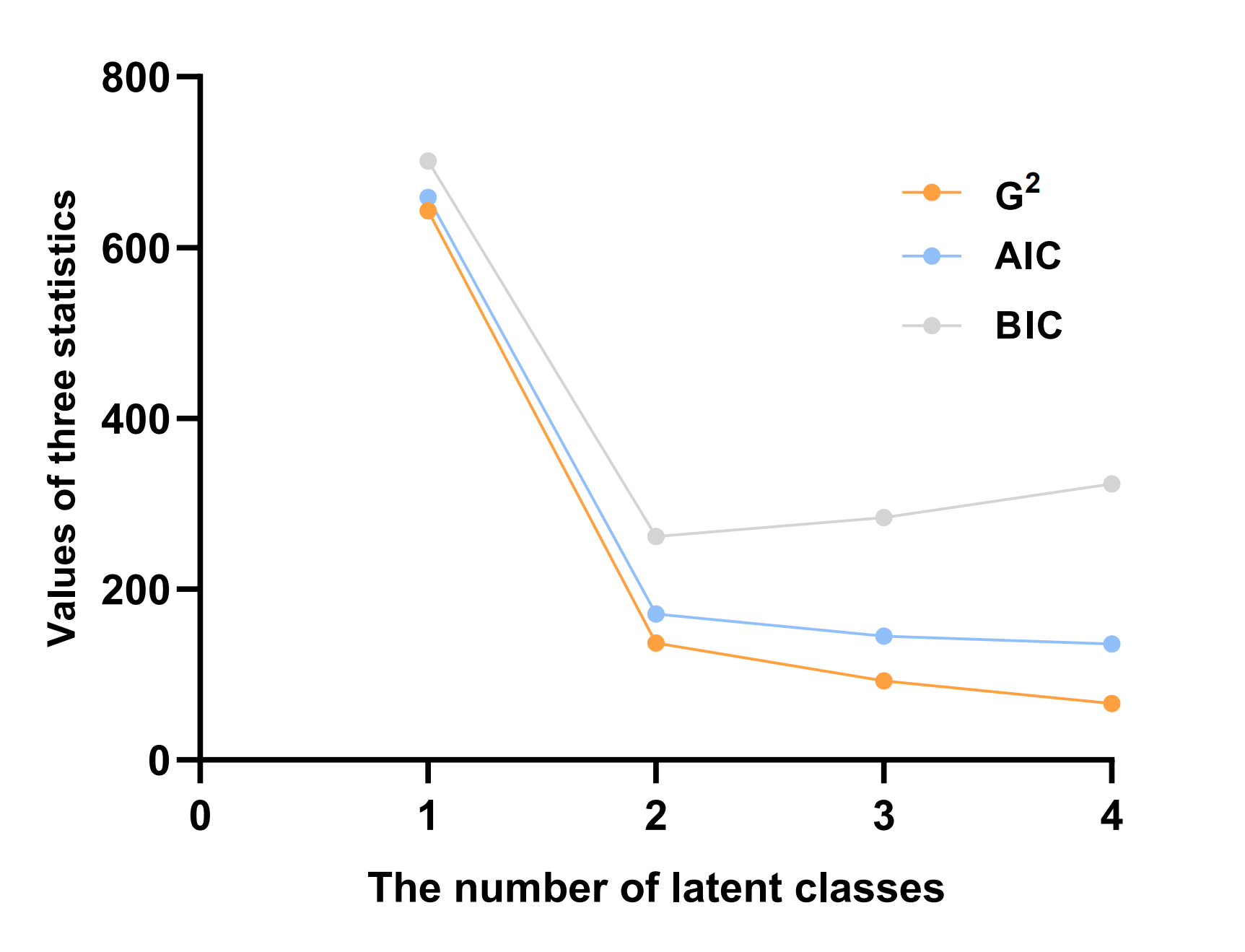

**Figure S2.** G2 statistics, AIC, and BIC in models with different numbers of latent classes in the US NHANES

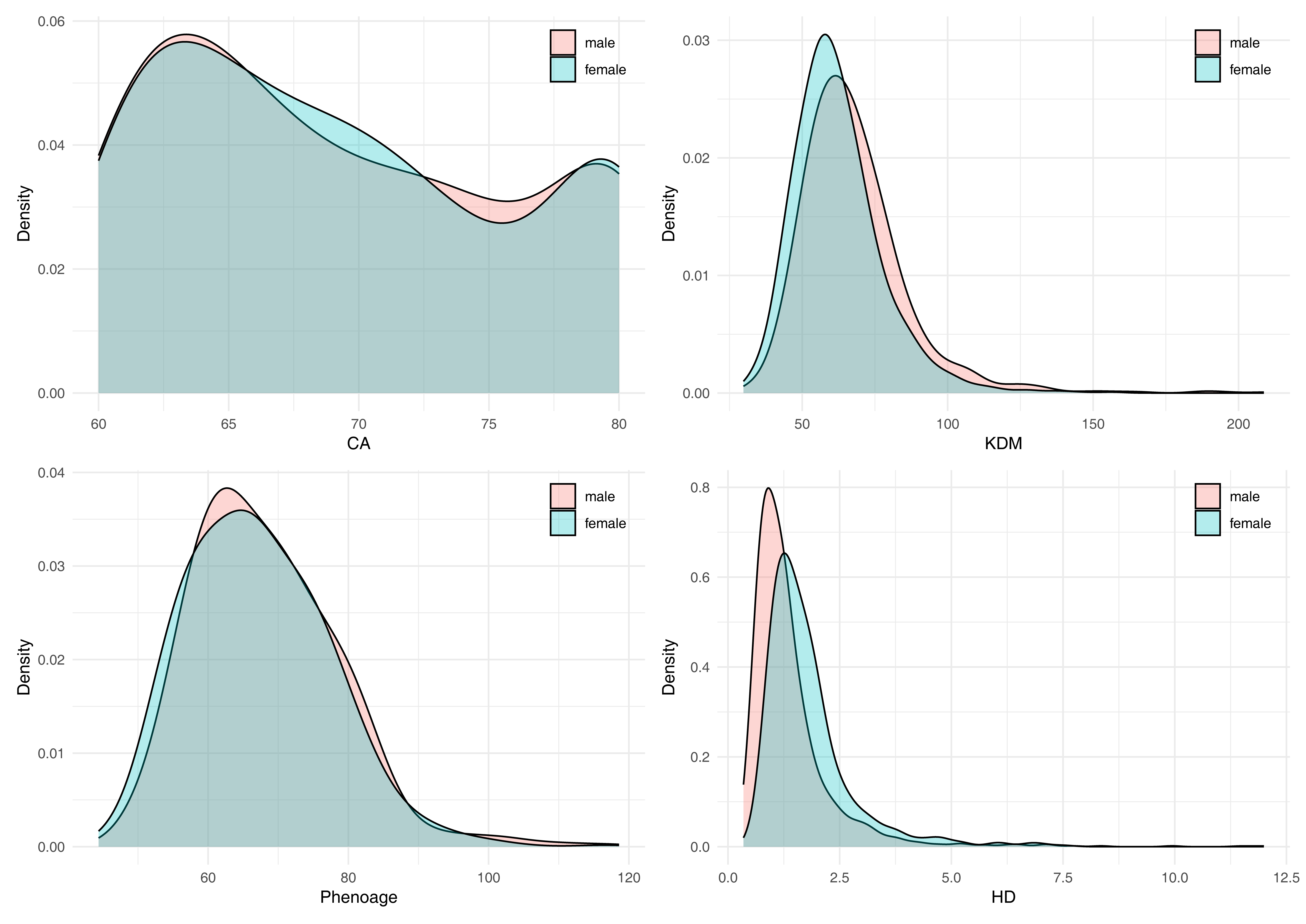

Figure S4. Distribution of chronological age and whole-body aging age measures.

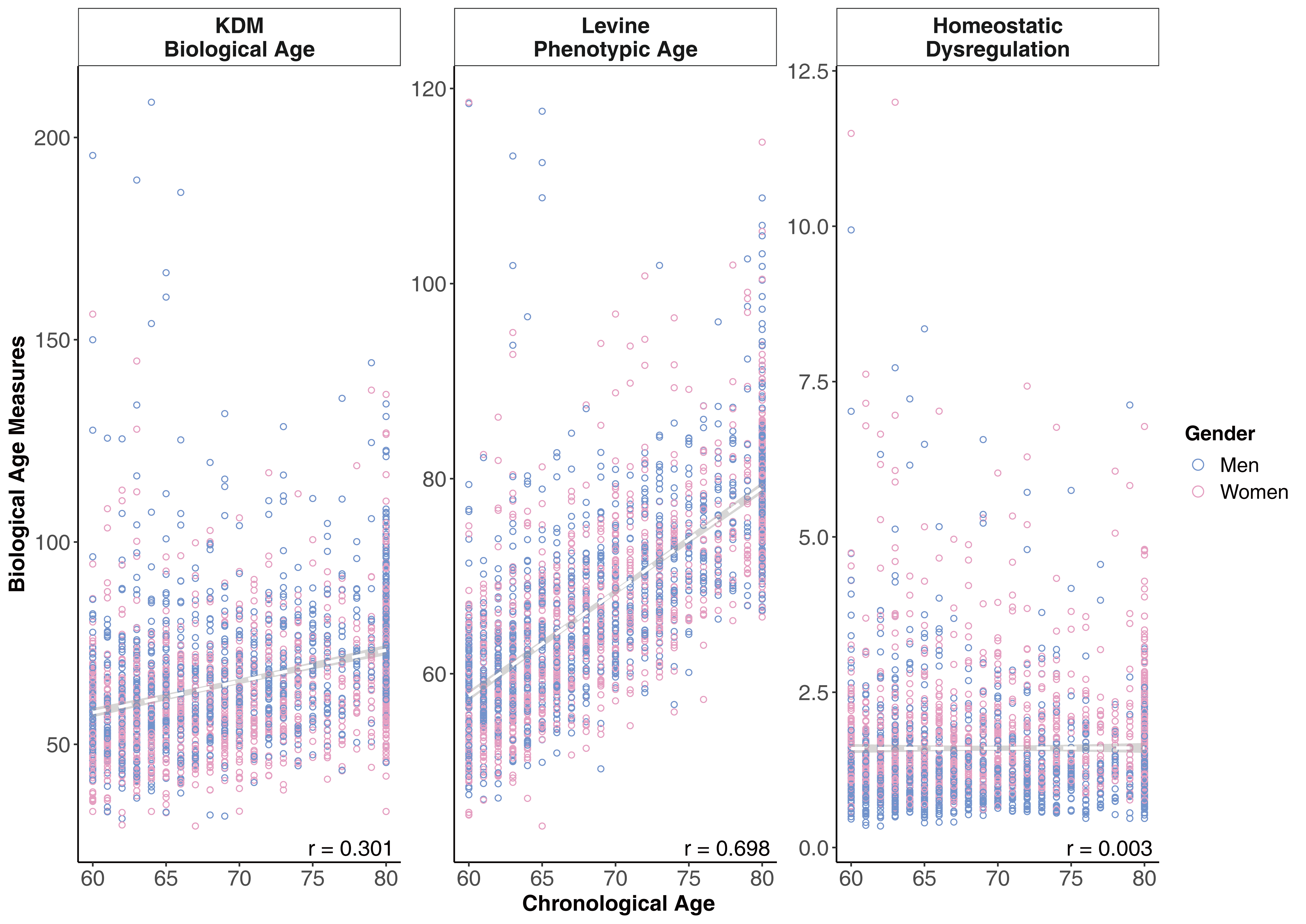

**Figure S4.** Scatter plots of whole-body aging measures and chronological age.
